# Formal dissent in CHMP opinions on new medicines, 2020-2025

**DOI:** 10.64898/2026.11.03.26367215

**Authors:** Maximilian Siebert, Ariadna Tibau

## Abstract

Disagreement within medicines regulation is rarely visible in the public record at the point an opinion is adopted. We examined divergent positions formally appended to opinions of the European Medicines Agency’s Committee for Medicinal Products for Human Use (CHMP) on new medicines from 2020 through 2025, both those opposing a positive opinion and those favoring authorization despite a negative opinion, describing how often they occurred, how large the dissenting minorities were, and which issues they raised. Divergent positions opposed 28 of 331 positive opinions (8.5%) and most often concerned efficacy and methodological limitations. Of 22 final negative opinions publicly available, 6 (27%) were accompanied by a divergent position favoring authorization. These records show when CHMP members disagreed about whether the evidence justified authorization. Indexing these disagreements and publishing vote outcomes with each opinion would make regulatory uncertainty visible and help clinicians, payers and health technology assessment bodies make better-informed decisions about treatment use and coverage.

## Background

Scientific disagreement is an inherent part of regulatory decision-making, but it is rarely visible in the final public record. Within the European Medicines Agency (EMA), members of the Committee for Medicinal Products for Human Use (CHMP) who do not support an adopted opinion may append a signed divergent position to the European Public Assessment Report, setting out the basis for their disagreement.^1^ These documents provide a distinctive window into disagreement within the body responsible for forming the EMA’s scientific opinion, rather than disagreement between a regulatory agency and an external advisory panel. Although individual divergent positions have been cited in previous regulatory-science analyses of the evidence supporting medicine approvals,^2,3^ their frequency, extent and stated grounds across recent opinions on new medicines have not been systematically characterized. Such records have been proposed as a standardized signal of regulatory confidence that should be indexed and reported alongside each opinion, particularly as the EU pharmaceutical reform and the health technology assessment regulation make the CHMP opinion more consequential for downstream decisions.^4^ We therefore examined divergent positions, including both dissents opposing positive opinions and dissents supporting authorization following a negative opinion.

## Methods

We reviewed CHMP opinions on new medicines adopted between 2020 and 2025 and identified those accompanied by at least one divergent position opposing a positive opinion. We separately identified negative opinions accompanied by a divergent position favoring authorization. We treated the vote margin as a characteristic of the opinion as a whole, and the grounds for dissent as characteristics of individual members’ positions.

We identified new medicines for human use that received a positive CHMP opinion between 1 January 2020 and 31 December 2025 from CHMP meeting highlights on the EMA website.^5,6^ Biosimilars, generics and hybrid medicines were excluded.

For each medicine, we reviewed the European Public Assessment Report (EPAR) and associated CHMP documents for divergent positions, defined as formal statements signed by one or more CHMP members recording disagreement with the adopted majority opinion.^1^ For each positive opinion accompanied by a divergent position, we extracted the opinion date, therapeutic area and indication, and regulatory characteristics (orphan designation, conditional marketing authorization, authorization under exceptional circumstances, accelerated assessment, PRIority Medicines (PRIME) eligibility, advanced therapy status). From the CHMP minutes,^5^ we extracted the number of members voting and the number voting in favor or against. For each divergent position, we recorded the number of signatories and coded the grounds for dissent into predefined, non-mutually exclusive categories: efficacy, safety, methodology (comprising study design, risk of bias, sample size and follow-up), quality of the evidence, proposed indication, and whether the position argued for a conditional rather than a full marketing authorization.

Efficacy was coded when a position questioned the magnitude, relevance or certainty of benefit, and safety when it cited specific adverse effects.

Negative opinions issued during the same period were examined separately to identify cases in which one or more CHMP members appended a divergent position favoring authorization. These cases were identified from CHMP meeting records and the corresponding EPAR documentation.^5^ For such refusal dissents, we extracted the same core variables as for positive opinions. All negative opinions on new medicines issued during the study period were enumerated from the CHMP meeting records. Negative opinions that were reversed on re-examination (n = 6) were analyzed as positive opinions and the remaining 22 were treated as final negative opinions.

Divergent positions were obtained from the EPARs, whereas voting results were extracted from the CHMP meeting records corresponding to the month in which the opinion was adopted. The same meeting records were used to identify negative opinions and determine whether they were accompanied by a divergent position. Where meeting documentation was not publicly available, we submitted four freedom-of-information requests to the EMA for the relevant CHMP meeting records. At the time of analysis (October 2026), the divergent-position document was unavailable for one opinion, and the voting record for one further opinion remained pending.

CHMP voting rules were considered when interpreting vote totals and signatories. Opinions are adopted by consensus where possible, otherwise an absolute majority of members eligible to vote is required, with a quorum of two thirds and if no absolute majority is obtained, the opinion is deemed negative. Members from the EEA-EFTA states (Iceland, Liechtenstein and Norway, which belong to the European Free Trade Association and the European Economic Area) may participate in the committee’s work and have a divergent position appended to its opinion, but they are not eligible to vote and their positions are excluded from the vote count.^1^ Accordingly, vote totals and the number of divergent-position signatories were treated as distinct variables. For Durveqtix, a separate divergent position adopted by the Committee for Advanced Therapies was not counted as an additional CHMP position, and Enerzair Breezhaler and Zimbus Breezhaler, which share a single dossier, were counted as one product.

We summarized counts and percentages of positions by category of dissenting rationale. Committee and dissent sizes were reported as medians with interquartile ranges, and the prevalence of dissent was estimated with a 95% Wilson confidence interval. Analyses were descriptive and no formal hypothesis testing or inferential modeling was performed.

## Results

Of 331 positive CHMP opinions (each followed by a European Commission marketing authorization) on new medicines, 28 (8.5%, 95% CI 6-12) were accompanied by at least one divergent position opposing the positive opinion. One product attracted two positions, giving 29 positions across 28 products. Positions were signed by a median of 3 members (IQR 2-5) and 7 were signed by a single member. The committee vote was recorded for 27 of the 28 products (median vote total 29, IQR 29-32). In these 27, the dissenting side accounted for a median of 3 recorded votes (IQR 2-6). It reached 15 of 32 votes (47%) in the two most contested opinions, for the anti-amyloid antibodies lecanemab and donanemab, where 16 members signed each divergent position (Figure 1a).

**Figure 1.**
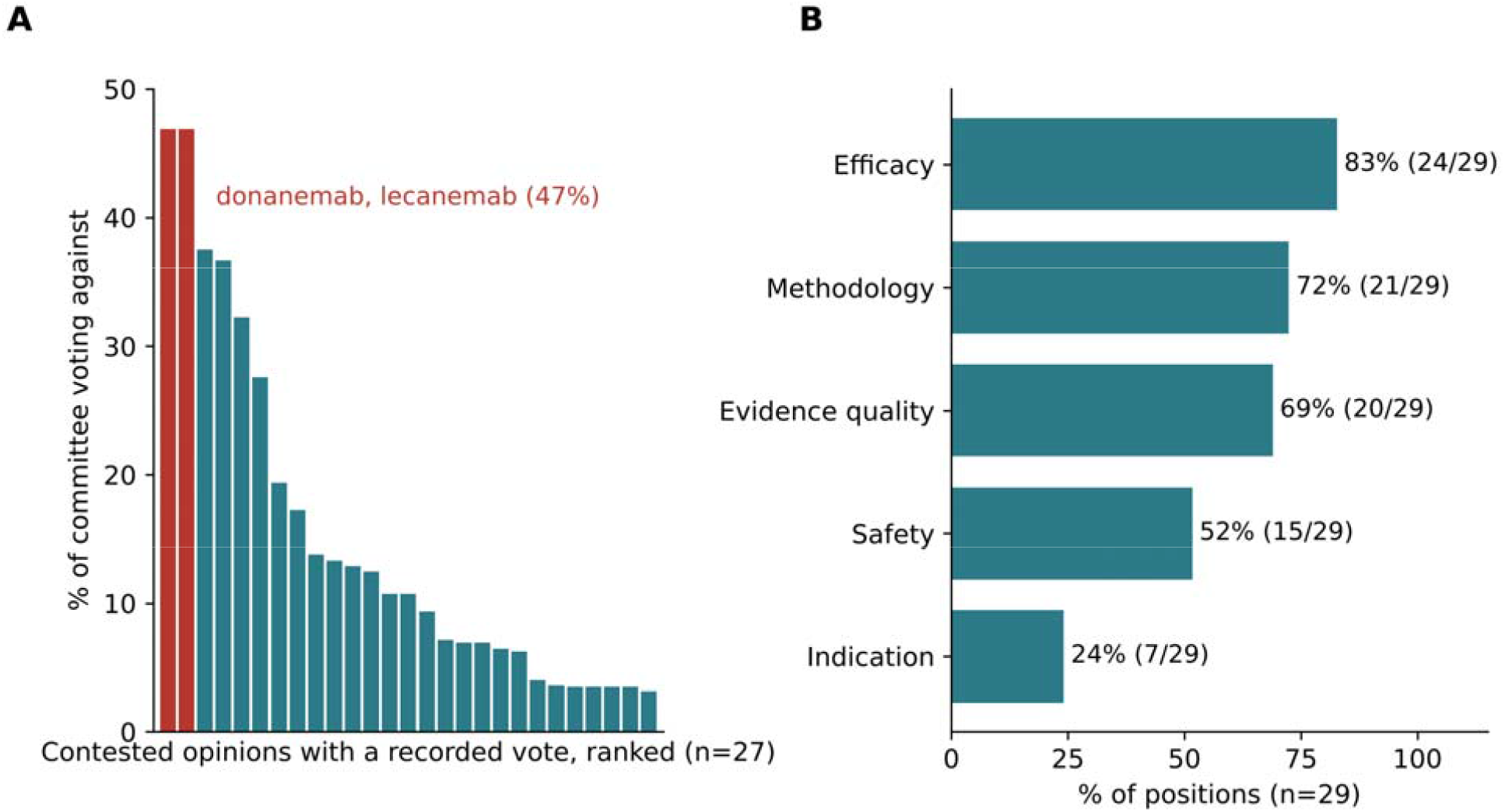
Size and grounds of CHMP divergent positions opposing a positive opinion, 2020-25. **a** Percentage of recorded votes cast against the adopted opinion (an opinion-level measure), for the 27 of 28 products with a recorded vote, ranked in descending order; one product carried two divergent positions, so the 29 positions arise from 28 products. The two highest are the anti-amyloid antibodies lecanemab and donanemab (15 of 32 votes against, 47%; red). **b** Grounds cited across the 29 divergent positions (position-level); positions could cite more than one ground, so percentages do not sum to 100. “Methodology” collapses study design, risk of bias, sample size and follow-up.

The 28 drugs with at least one divergent position spanned nine therapeutic areas. Oncology indications (i.e. solid tumors) accounted for half (14; 50%), followed by neurology (4; 14%), non-malignant hematology and hemostasis indications (2; 7%), pneumology/allergology (2; 7%) and dermatology (2; 7%). Most were small molecules (17; 61%), and more than half (15; 54%) had at least one special regulatory pathway or designation, most often conditional marketing authorization (8; 29%) or orphan designation (7; 25%) (Table 1). Across the 29 positions, efficacy was contested in 24 (83%), methodological limitations in 21 (72%), the quality of the evidence in 20 (69%), safety in 15 (52%), and the proposed indication in 7 (24%) (Figure 1b). As these were not mutually exclusive, positions frequently cited more than one ground. In four positions, members argued explicitly that the premarketing evidence supported only a conditional and not a full marketing authorization. In their view, the residual uncertainty was considered compatible with approval only if it was addressed through specific obligations, the binding post-authorization evidence requirements attached to a conditional marketing authorization. In three of these, full approval was nonetheless granted. These concerns correspond to evidentiary features that the approvals literature has shown to be common at the EMA and elsewhere, including single-arm trials, unvalidated surrogate endpoints, reliance on a single pivotal study and pivotal trials that missed their prespecified primary endpoints.^7,8^

**Table 1.** Characteristics of medicines authorized despite a divergent position (n = 28)

| **Characteristic** | **n (%)** |
|---|---|
| **Therapeutic area** |  |
| Oncology (solid tumors) | 14 (50) |
| Neurology | 4 (14) |
| Non-malignant hematology/hemostasis | 2 (7) |
| Pneumology/allergology | 2 (7) |
| Dermatology | 2 (7) |
| Other* | 4 (14) |
| **Year of opinion** |  |
| 2020 | 5 (18) |
| 2021 | 5 (18) |
| 2022 | 2 (7) |
| 2023 | 3 (11) |
| 2024 | 8 (29) |
| 2025 | 5 (18) |
| **Type of medicinal product** |  |
| Small molecule | 17 (61) |
| Biological | 10 (36) |
| Advanced therapy medicinal product | 1 (4) |
| **Regulatory programs and designations** |  |
| Conditional marketing authorization | 8 (29) |
| Authorization under exceptional circumstances | 3 (11) |
| Accelerated assessment | 0 (0) |
| PRIority Medicines (PRIME) | 3 (11) |
| Orphan designation | 7 (25) |
| Any special regulatory pathway or designation<sup>+</sup> | 15 (54) |

\*One each: ophthalmology, immunological/inflammatory, endocrinology and diagnostic agent
<sup>+</sup>Conditional marketing authorization, marketing authorization under exceptional circumstances, accelerated assessment, PRIME and orphan designation.

Formal dissent was uncommon but occasionally substantial. In the re-examination votes for lecanemab and donanemab, 15 of the 32 votes recorded in each case opposed the adopted opinion. However, this degree of division is only partially visible outside the assessment reports. An account of these assessments by EMA staff and CHMP members reported that both re-examination opinions were adopted by majority and summarized the minority view that the effects were too small to outweigh the risk of amyloid-related imaging abnormalities.^9^

Dissent also accompanied negative opinions. Of the 22 final negative opinions, 6 (27%) were accompanied by a divergent position favoring authorization. Where reported, the number of signatories ranged from 2 to 11 (median 6), and the majority opposing authorization ranged from 72% (21 of 29 votes) to 93% (27 of 29). In all five positions with available text, the minority considered the benefit-risk balance positive. Moreover, all five addressed efficacy and safety, and three acknowledged limitations in the quality of the evidence but judged them tolerable. Rather than disputing the majority’s interpretation of the evidence, these positions generally argued that evidentiary standards should reflect disease severity and the absence of alternatives. Three products relied on surrogate or non-patient-centered endpoints, and three proposed addressing residual uncertainty after approval through conditional authorization and specific obligations. The conditional pathway was therefore invoked in both directions, both to argue that evidence judged sufficient warranted only conditional approval and to argue that evidence judged insufficient warranted conditional approval rather than refusal. The largest minority concerned pegcetacoplan (Syfovre) for geographic atrophy secondary to age-related macular degeneration. Eleven members signed a divergent position, although the committee vote was not available. The signatories considered slowed lesion growth clinically meaningful in a slowly progressing disease with no approved treatment in the EU, whereas the majority judged that it had not been shown to translate into functional benefit for patients.

## Discussion

Formal dissent accompanied around 9% of positive CHMP opinions on new medicines between 2020 and 2025. Dissent against positive opinions most often questioned efficacy and the methodological basis of the evidence, whereas dissent against refusals generally accepted the evidentiary limitations but judged them tolerable given disease severity and the absence of alternatives.

CHMP vote splits, however, are not directly comparable with US Food and Drug Administration (FDA) advisory-committee votes, because the two bodies occupy different positions in the decision-making process. FDA advisory committees are external to the agency’s decision-making chain and their votes are recommendations and may be followed or rejected, as illustrated by aducanumab.^10,11^ CHMP divergent positions instead document disagreement among members of the committee that adopts the EMA scientific opinion. The European Commission then makes the legally binding marketing-authorization decision. Disagreement within the FDA itself is documented in approval packages rather than through votes: among 174 novel therapeutics approved in 2011-2015, nearly a quarter had at least one documented disagreement among reviewers or leadership and 7% included a disagreement about whether to approve.^12^ CHMP vote splits therefore measure division within the opinion-forming committee, whereas FDA advisory-committee votes measure advice supplied to the regulator.^13^

Despite our findings, vote distributions and divergent positions should be interpreted narrowly. A vote split quantifies disagreement over the committee’s adopted position, but it does not identify why members disagreed or provide a calibrated measure of evidentiary uncertainty. Members may assign different weight to clinical relevance, safety, the target population, benefit-risk trade-offs, or the regulatory mechanism used to manage remaining uncertainty. Read together with the accompanying statements, however, vote distributions show both the extent of disagreement and the specific issues on which the minority departed from the majority.

Our analysis has limitations. It is confined to dissent that members chose to record formally, over a relatively short window, refusals later reversed on re-examination were analyzed as positive opinions rather than as refusals, and the text of one refusal dissent was unavailable. Furthermore, we did not compute within-therapeutic-area rates of dissent. We also cannot determine from these documents what produced a disagreement or whether factors external to the evidence influenced individual judgments, questions considered elsewhere.^14^

These data also suggest that divergent positions could be tracked over time as an indicator of regulatory uncertainty and linked to what happens to medicines after approval. Linking each position to the corresponding vote and regulatory pathway would permit analyses of whether concerns raised at the time of the opinion were followed by confirmatory evidence, indication changes, additional risk-management measures, or other post-authorization action. The latter would make formal dissent usable for studying the evolution of regulatory evidence rather than treating it only as a feature of individual assessment reports. Indexing divergent positions and publishing vote outcomes with each opinion would make such linkage routine.^4^

## Data Availability

Data are available on request

## Declarations

## Author contributions

M.S. conceived and designed the study, collected and analyzed the data, and drafted the manuscript. A.T. contributed to study design, data interpretation, and critical revision. Both authors approved the final version.

